# Community vulnerability to epidemics in Nepal: A high-resolution spatial assessment amidst COVID-19 pandemic

**DOI:** 10.1101/2020.07.01.20144113

**Authors:** Laxman Khanal, Binod Kumar Paudel, Bipin Kumar Acharya

## Abstract

The coronavirus disease 19 (COVID-19), the biggest health problem at present, doesn’t have uniform transmission and severity among the countries and communities therein. Knowledge of community vulnerability to the disease would facilitate interventions aimed at transmission control by the efficient deployment of available limited resources. Therefore, we assessed spatial variations and heterogeneity of disease vulnerability among the population in 753 municipal units of Nepal. We collected geospatial indicators representing the domain of socioeconomic inequalities, population dynamics, heterogeneity in accessibility and the information related to underlying health condition which potentially affect the severity of COVID-19 transmission. Those indicators were assembled to create three vulnerability indices using Geographic Information System (GIS); Social Vulnerability Index (SVI), Epidemiological Vulnerability Index (EVI) and a composite of the two- Social and Epidemiological Vulnerability Index (SEVI). The indicators were scaled to a common measurement scale and spatially overlaid via equally weighted arithmetic mean. The indices were classified into five level of risk and the municipal units and the population within vulnerabilities classes were quantified and visualized in the map. The index output indicated high vulnerability to epidemics in megacities like Kathmandu, Pokhara, Bharatpur, etc.; developing cities especially in the Province No 2; and, municipal units of Karnali and Sudoorpashchim provinces. Additionally, some other municipalities such as Dhulikhel, Beshishahar, Tansen etc. which have a higher prevalence of pulmonary and cardiovascular disorders are highly vulnerable. The SVI indicated that 174 municipal units and 41.5% population is highly vulnerable. The EVI identified 55 municipal units and 40.7% of the total population of the country highly vulnerable to COVID-19. The SEVI accounted that disease vulnerability is high in 105 municipal units and 40% population of Nepal. The vulnerability indices created are means for different tiers of the existing government in federal system of Nepal for prioritization and improved planning for disease intervention especially in highly vulnerable municipal units where the COVID-19 transmission could have high severity.

## 1 Introduction

Coronavirus disease 2019 (COVID-19) is a zoonotic disease caused by an RNA virus known as severe acute respiratory syndrome coronavirus 2 (SARS-CoV-2) reported from Wuhan, China in December 2019 (Huang et al. 2020). The World Health Organization (WHO) has declared the disease (COVID-19) a Public Health Emergency of International Concern (PHEIC) on 31^st^ January 2020 (WHO 2020a). The COVID-19 pandemic, as of 25^th^ June 2020, has spread exponentially to 215 countries, areas and territories, infecting 9,296,202 people and causing over 479,133 deaths (WHO 2020b). The mortality rate of the disease has been 5.15 per thousand infections. The intensity of its spread and the rate of mortality is not even among the spatial extents like continents, countries and geographical areas such as counties; communities, age-groups and sexes (Baud et al. 2020, Wadhera et al. 2020, WHO 2020b). Higher rate of transmission and mortality have been recorded from the areas of dense settlement and heavy flow of people. About 80% of people with COVID-19 have mild or asymptomatic disease, therefore, symptom-based control is unlikely to be sufficient (Anderson et al. 2020, WHO 2020a). The basic reproduction number (R_0_) of COVID-19, i.e. the mean number of secondary cases generated by one primary case when the population is largely susceptible to infection, had a value of about 2.5 in the earlier days of outbreak (Liu et al. 2020) that has substantially been lowered with the impose of social distancing measures (Rothan & Byrareddy 2020). The greater the reduction in transmission, the longer and flatter will be the epidemic curve; for which early self-isolation and social distancing are the keys. Despite rigorous global efforts, COVID-19 is continuously spreading across the world causing significant illness and death (WHO 2020b). No vaccine or effective antiviral drug is likely to be available soon as the process will take time (Anderson et al. 2020).

Severity of a pandemic is not uniform in all societies (Wadhera et al. 2020). Social vulnerability theory posits that “societal inequalities exist based on class, race, ethnicity, gender, age, health, abilities, etc.” (Cannon 1994). Differences in socioeconomic factors impose different degree of impacts from hazards (Mwarumba 2017). Vulnerability includes exposure to detrimental environmental or social strains, susceptibility to those strains, and the capacity to adapt (Adger 2006). Being more specific to a disease, vulnerability is the elevated exposure to infection; increased susceptibility to the disease including its complications, hospitalizations, and death; and lack of access to health care (Frankel 2011). People living with smallest number of choices and forced lives from poverty, gender and ethnic discrimination, physical disability, limited employment opportunities, and other forms of domination are the most vulnerable to such disasters (Cannon 1994). Severity indices of the community to the disease could explain possible relationships underlying the temporal and spatial aspects of its diffusion.

The severity of COVID-19 is higher in areas of higher population density (Coccia 2020, Rocklöv & Sjödin 2020, Wadhera et al. 2020) and their movement (Tuite et al. 2020, You et al. 2020). Higher population density lowers the efficacy of social distancing and adds complications in the contact tracing of the COVID-19 patients that ultimately increases the R_0_ (Anderson et al. 2020, Rocklöv & Sjödin 2020). The disease incidence in children seems to be lower than in the rest of the population (Sominsky et al. 2020, Wu & McGoogan 2020), so is the disease accounted mortality rate (Dong et al. 2020). Pre-existing chronic medical conditions such as chronic obstructive pulmonary disease (COPD), cardiovascular diseases such as high blood pressure, diabetes etc. may predispose adults to more severe outcomes of the disease (Fang et al. 2020, Yang et al. 2020, Zheng et al. 2020). The innate immune mechanism is less efficient in elderly people that makes them more vulnerable to severe outcomes of infectious diseases (Boe et al. 2017). Communities having dense population dominated by the elderly people with compromised immunity presumably have higher vulnerability to the COVID-19 disease.

In response to the COVID-19 pandemic, most of the countries across the globe have imposed lockdown, due to which a large number of people have lost their job. Such people rush to their country and hometown, legally or illegally, without proper precautions. The movement of large groups of unprotected, non-immune, physically weakened and possibly infected people between different zones can increase the vulnerability of migrants and their host community to the pandemic (Bates et al. 2004). Poverty is one of the major determinants of the disease vulnerability as poor people have limitations in resource mobility and disease treatment. Hence, poverty is known as the “carrier status” of disease (Semenza & Giesecke 2008). Delays in diagnosis and treatment of epidemics are associated with worse transmission, morbidity and mortality (Bates et al. 2004). Additionally, poverty associated illiteracy leads to disproportionate misinformation and miscommunication among individuals with less access to information channels, who fail to maintain necessary precautions against the disease. Undernutrition and lack of sanitation can worsen the situation of the pandemic by boosted diffusion and higher mortality. Health facilities are not adequate and accessible to all people in developing countries. Inequalities in geographic accessibility to healthcare even in developed countries like the United States have been documented to cause negative health outcomes for epidemics (Mollalo et al. 2020). Inability to access health care is a major obstacle to initiation of early treatment and prevent the transmission dynamics. Timely diagnosis, isolation and treatment is crucial for the control of pandemics like COVID-19 (Ji et al. 2020). Because almost all of the medical facilities meeting basic requirements for COVID-19 identification and treatment are in the city areas, travel time to hospitals and cities can be one of the important determinants of the vulnerability.

Large number of literatures have surged in the short period of time after COVID-19 outbreak, however, most of them are from the medical sciences using clinical and serological data. For the proper management and control of such pandemics it is equally important to know the transmission dynamics and identify the vulnerable group of people. Geospatial analysis using socioeconomic, demographic and geophysical data would be helpful to restrain the disease. Geographic information system (GIS) is an essential tool to examine the spatial distribution of infectious diseases (Acharya et al. 2018, Kang et al. 2020). However, a limited number of GIS-based studies have been published since the initial outbreak of COVID-19 (Kamel Boulos & Geraghty 2020, Kang et al. 2020, Macharia et al. 2020, Mollalo et al. 2020, Sarwar et al. 2020).

Assessment of the disease risk level is important for preparedness and response. It is vital to explain the determining factors of the transmission dynamics of this highly infectious disease for designing strategies to control diffusion, empowering health policy with economic, social and environmental interventions (Coccia 2020). However, real risk of infection and death depends on a number of factors including domain of population structure, socioeconomic heterogeneity and underlying health condition of population. By the date of 25^th^ June 2020, COVID-19 has accounted a total of 11,162 infections and 26 deaths from 76 of the 77 districts within Nepal territory (MoHP 2020). Most of those cases of the disease in Nepal are among mass return migrants from India, China and other areas, except few suspected community level transmission cases in Birgung, Nepalgunj and Udaypur (Tharu et al. 2020). If the community level transmission of COVID-19 or other epidemics starts, it is important to identify the most vulnerable communities and areas such that available limited resources could be efficiently and timely deployed in those priority zones. This knowledge would facilitate interventions aimed at transmission control and patient care, minimizing the collective and individual burden of the pandemic. Therefore, we aimed to explore-i) demographic and health resource accessibility inequities; ii) socioeconomic inequities; iii) disease prevalence inequities; and, iii) severity indices to epidemics among 753 municipal units of Nepal. We used demographic, socioeconomic and spatial data, and GIS tools to identify the Social Vulnerability Index (SVI), Epidemiological Vulnerability Index (EVI) and a composite of the two, the Social-Epidemiological Vulnerability Index (SEVI).

## 2 Methods

### 2.1 Study area

This study was conducted covering entire 753 municipal units in Nepal. The municipal units include four categories: six metropolitan cities, 11 sub-metropolitan cities, 276 municipalities and 460 rural municipalities. These units are third tire of government in the federal structure of Nepal. Administratively these municipal units come under third hierarchy after the province (n=7) and district (n=77) units in Nepal.

### 2.2 Data collection and processing

Based on the previous findings and availability of freely accessible data sources, we collected 14 relevant data layers for this assessment covering four broad domains of vulnerability-socio economic inequalities (Mollalo et al. 2020), population dynamics (Wadhera et al. 2020), access to health services (Rader et al. 2020,2020, Smith & Judd 2020, Wadhera et al. 2020) and underlying health condition such as prevalence of hypertension, diabetes, asthma and COPD.

#### Socioeconomic inequalities data

Proportion of household with no toilet facility, proportion of households with access to improved water sources, literacy and prevalence of stunting among children were five indicators related with socioeconomic inequalities which were collected as gridded raster layers from demographic and health survey (DHS) 2016 (https://www.dhsprogram.com). These raster surfaces were generated using geostatistical modeling based DHS indicators and several relevant environmental covariates (Mayala et al. 2019). In addition, poverty status raster layer generated using geostatistical method-based household wealth quantile data of DHS and relevant environmental covariates was also retrieved from the worldpop geoportal (https://www.worldpop.org/). Then, mean values of these layers were extracted using the zonal statistics function of the spatial analyst tool of ArcGIS 10.5.

#### Demographic and accessibility data

Population density and elderly population of age above 60 years; two demographic indicators used in the study were retrieved for the year of 2020 from the worldpop geoportal (https://www.worldpop.org/) in gridded format. The mean value of these layers was summarized in municipal unit using zonal statistics tool of ArcGIS 10.5. To normalize the distribution of population, we took log population density instead of direct population density. Proportion of elderly population were computed by dividing elderly population with total population and expressed in percentage. For the accessibility, we used two indicators; accessibility to urban center and accessibility to health facilities. For urban accessibility, travel time to nearest urban center with population higher than 50,000 were retrieved from Weiss et al. (2018) and extracted in the extent of Nepal while accessibility to health facilities was computed by ourselves. We collected hospital location data from open access Humanitarian Data Exchange Portal (https://data.humdata.org/dataset/nepal-health-facilities-cod), landcover map of 2010 from ICIMOD geoportal (Uddin et al. 2015), road from open street map (https://www.openstreetmap.org/) and SRTM DEM from consortium for spatial information geoportal (http://srtm.csi.cgiar.org/srtmdata/) for this purpose. We used AccessMod version 5.0 (Ray & Ebener 2008) to process these datasets and compute the travel time. Median travel time to nearest health facilities was calculated as a cost distance depending on specific cost values for different land cover properties and considering topographical barriers. The median value of travel time raster and urban accessibility raster were summarized in local unit level using the zonal statistics tools of ArcGIS 10.5.

#### Epidemiological data

Reported number of cases for four major non-communicable diseases: hypertension, diabetes, asthma and COPD in Out Patient Department (OPD) services of different hospitals aggregated in municipal unit were downloaded from Health Management Information System (HMIS) web portal (https://dohs.gov.np/ihims-raw-data/) first. Similarly, municipal unit wise population of Nepal was also retrieved from Central Bureau of Statistics (https://cbs.gov.np/population-of-753-local-unit/). Then, both the datasets were linked to 753 municipal units in Arc GIS. Using the GeoDa software (Anselin et al. 2006), prevalence rate per thousand of population were computed for all four diseases. Considering possible instability prevalence rate due to small population, empirical Bayes smoothing was used based on K-nearest neighbored method (Pringle 1996). The indicators used and their nature, data format, resolution and sources are presented in the Table 1. The methodological workflow for this study is summarized in graphical form in the Figure 2.

**Table 1.** Demographic and socioeconomic inequality data used in the study and their sources

| Indicators | Description of Indicators | Data Format | Resolution | Year/Source |
| --- | --- | --- | --- | --- |
| **Toilet facility** | Proportion of households with no toilet facility | Raster | 1*1 km | 2016<sup>a</sup> |
| **Dirking water facility** | Proportion of households with access to improved water sources | Raster | 1*1 km | 2016<sup>a</sup> |
| **Malnutrition** | Prevalence of stunting among children | Raster | 1*1 km | 2016<sup>a</sup> |
| **Literacy** | Percentage of people are literate | Raster | 1*1 km | 2016<sup>a</sup> |
| **Poverty** | Wealth Index | Raster | 1*1 km | 2016<sup>b</sup> |
| **Log Population density** | Total population per unit area | Raster | 1*1 km | 2016<sup>b</sup> |
| **Elderly population** | Percentage of the population aged 60+ | Raster | 1*1 km | 2016<sup>b</sup> |
| **Access to hospitals** | Median travel time to nearest hospital | Raster | 100*100 m | 2016<sup>c</sup> |
| **Access to urban areas** | Travel time to the nearest urban center with at ≥ 5000 people | Raster | 1*1 km | 2015<sup>d</sup> |
| **Hypertension** | Prevalence of hypertension per 1000 | Aggregated Count | municipal unit | 2018<sup>e</sup> |
| **COPD** | Prevalence of COPD per 1000 | Aggregated Count | municipal unit | 2018<sup>e</sup> |
| **Diabetes** | Prevalence of diabetes per 100 | Aggregated Count | municipal unit | 2018<sup>e</sup> |
| **Bronchial Asthma** | Prevalence of asthma per 1000 | Aggregated Count | municipal unit | 2018<sup>e</sup> |
| **Notes:** a: https://www.dhsprogram.com/; b: https://www.worldpop.org/; c: Our own computation as explained above; d: https://malariaatlas.org/research-project/accessibility-to-cities/; and e: https://dohs.gov.np/ihims-raw-data |  |  |  |  |

**Figure 1.**
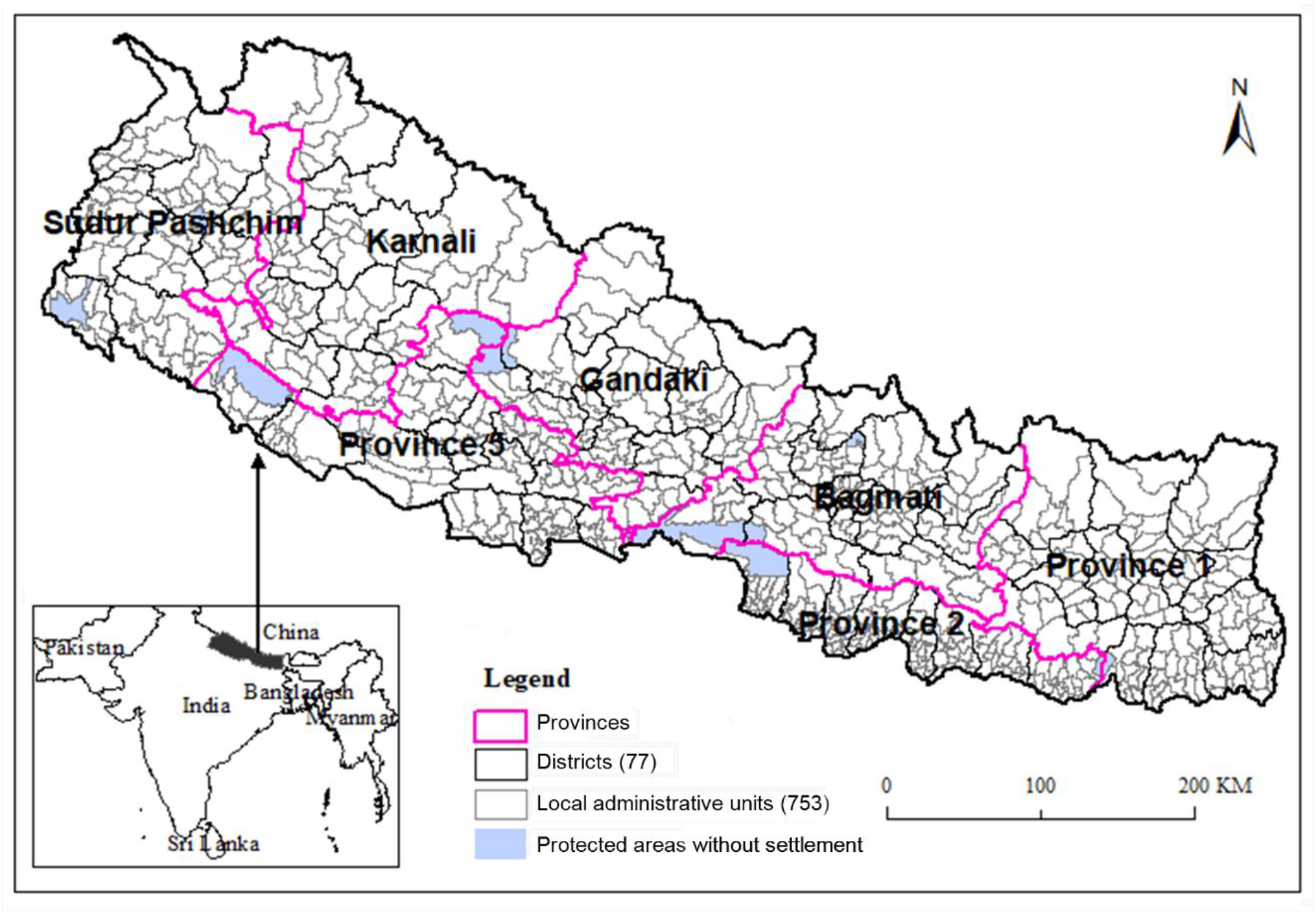
Map of Nepal showing 753 municipal units under 77 districts and seven provinces.

**Figure 2.**
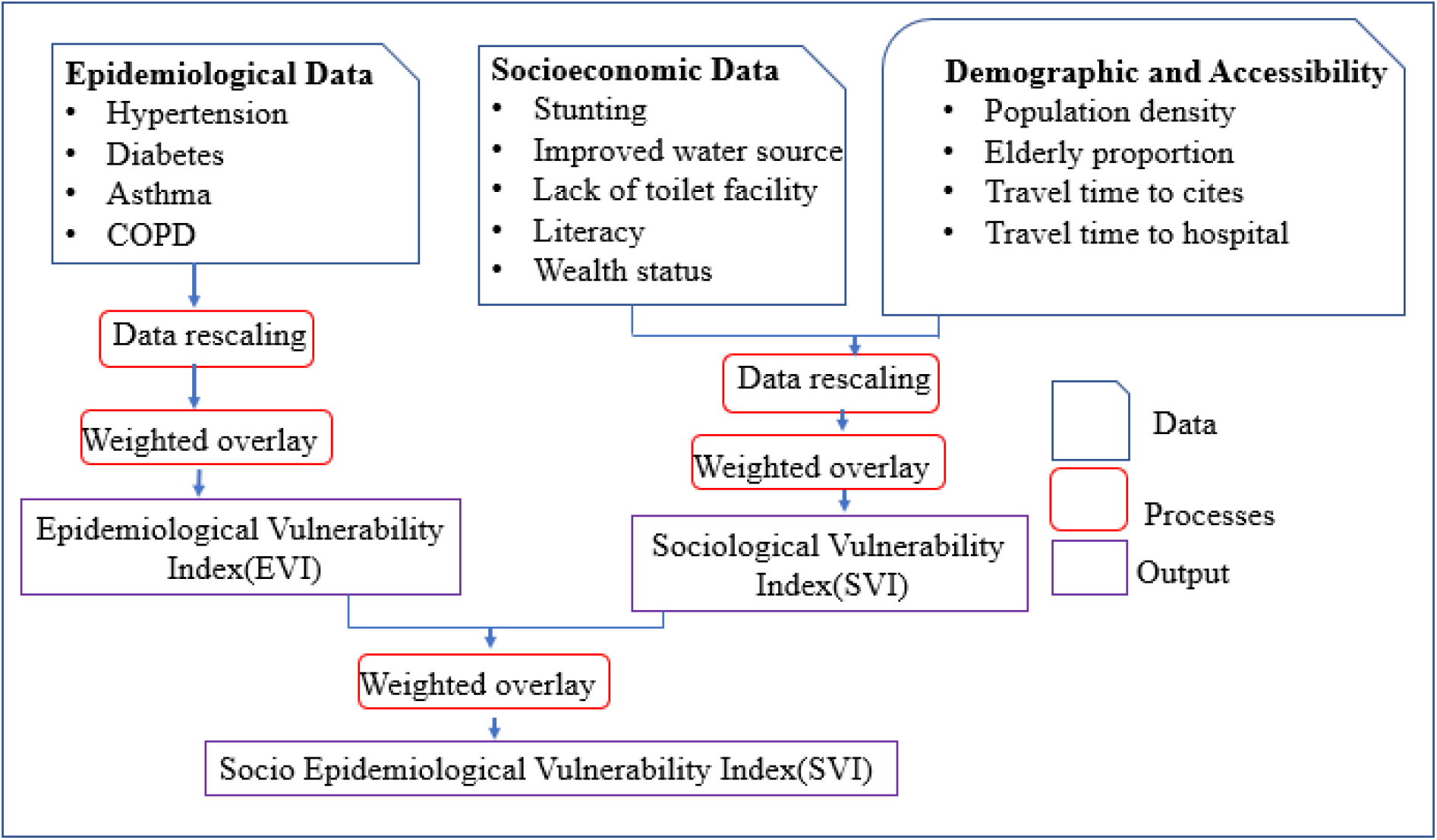
Methodological workflow for identification of community vulnerability

### 2.3 Vulnerability indices computation

As these potential indicators have different scales with different minima and maxima values, in the second step, all indicators were rescaled to a common scale ranging their values from 0 with least vulnerable to 100 most vulnerable to make values comparable using the equation (1). During this normalization process the direction of the indicators were also adjusted according to whether the indicator contributes positively or negatively.

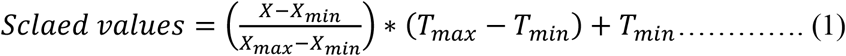

Where X denotes the value to be scaled; X_max_, X_min_ maximum and minimum values in the original range; and T_max_ and T_min_ maximum (100) and minimum (0) values to be scaled respectively.

We constructed three vulnerability indices: Social Vulnerability Index (SVI), Epidemiological Vulnerability Index (EVI) and a composite of the two, the Social-Epidemiological Vulnerability Index (SEVI) following the approach of (Macharia et al. 2020). We chose equally weighted arithmetic mean (Sullivan & Meigh 2006) for overlaying these indicators. However, in a reality, different indicators may affect vulnerability differently, we chose this method due to limited knowledge on relative contribution of these variables on community vulnerability of COVID-19. For the SVI, nine factors representing the socioeconomic inequalities, demographic dynamics and heterogeneity in accessibility were considered while for the EVI, prevalence of four chronic diseases were considered. The actual vulnerability was computed by integrating the both.

Finally, all three indices were visualized in ArcGIS environment by grouping them into five levels of risk: very low, low, moderate, high and very high, based on the natural junk classification method (Jenks 1977). The advantage of this classification method is it identifies “natural” groups within the data by reducing the variance within classes and maximizing the variance between classes (Jenks 1977). While many vulnerability assessment approaches provide tabular outputs, visualization is an important part of vulnerability assessment because it allows users to gain a better grasp of the spatial distribution of high or low vulnerability. End-users, and especially those without a technical background, can easily see and interpret the index output in this format compared to other formats (Dickin et al. 2013).

## 3 Results

### 3.1 Demographic and accessibility inequities

Among 753 municipal units of Nepal, municipalities including mega cities like Biratnagar, Bharatpur, Kathmandu, Pokhara, Butwal, Nepalgunj, Dhangadhi, etc. have high population density (above 809, up to 63652 individuals per square kilometer), much higher than the average national value (203 per square kilometer) (Fig. 3). Age wise structure of the population reveals that majority of municipal units in Bagmati and Gandaki provinces have higher proportion of elderly population (above 9.4% of the total population aged over 60 years) than rest of the municipalities and rural municipalities. Districts from the highland areas of Province No. 1, Karnali province and Sudoorpashchim province have highest required travel time to the hospital and cities.

**Figure 3.**
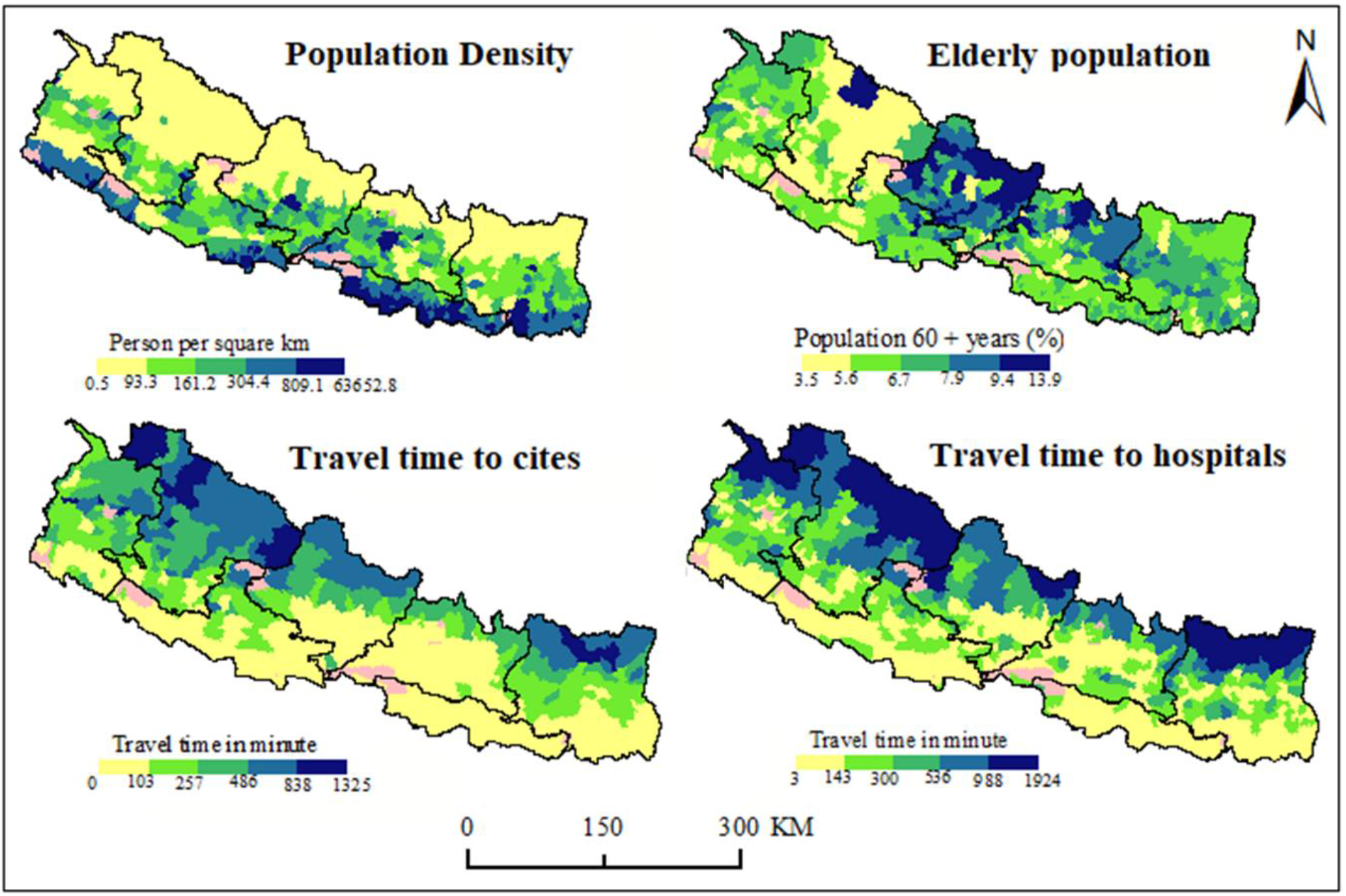
Spatial distribution of demographic and accessibility factors in municipal units of Nepal.

### 3.2 Socioeconomic inequities

Literacy rate is low in Rukum (east) district of Province No 5, sparsely populated some Himalayan districts, and densely populated districts of the Province No. 2 (Fig. 4). Both the poverty and stunting indices are above the national average values in Himalayan areas, especially in Karnali province and Sudoorpashchim province. Drinking water facility is poor in some pocket areas including Rukum (east) district. Coupled with high population density and illiteracy, the Province No. 2 with almost 2/3^rd^ population without the toilet facility has the worst situation of sanitation facilities.

**Figure 4.**
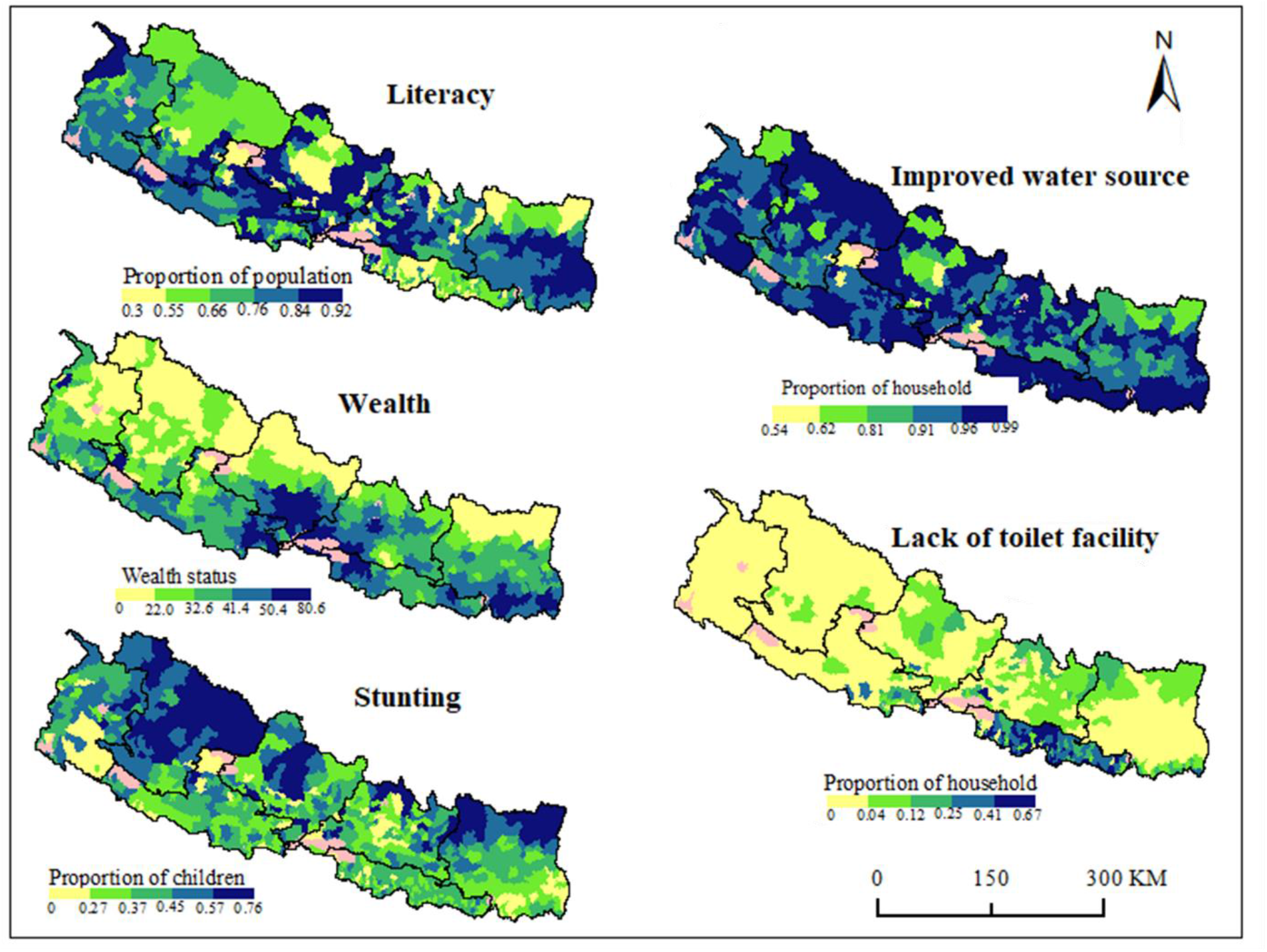
Spatial distribution of the socioeconomic inequalities among the municipal units of Nepal

### 3.3 Disease prevalence inequities

Prevalence of the diseases among the population is not uniform among the municipal units of Nepal (Fig. 5). The population in growing cities like Dhulikhel, Tansen etc. have the highest hypertension prevalence (above 200 individuals per 10,000). Asthma is more prevalent (35-57 per 10,000 individuals) among the districts of Karnali province (Kalimati, Palata, Kharpunath, Sarkegad Thulibheri, etc. RMs), Sudoorpashchim province (Amargadhi, Ajaymeru, Nawadurga from Dadeldhura district; Jayaprithivi RM from Bajhang; etc.), and some cities like Beshishahar in Lamjung, Dhorpatan in Baglung, etc. The chronic obstructive pulmonary disease (COPD) has higher prevalence among the population of Dhulikhel (121.9 per 10,000), Amargadhi (77 per 10,000), Sarkegadh (65 per 10,000), Tansen (62 per 10,000), Bhimeswor (61 per 10,000), etc. Diabetes is more prevalent in city areas such as Dhulikhel (122.4 per 10,000), Tansen, Siddharthnagar, Bharatpur, Biratnagar, Pokhara, Birtamod (50.4 per 10,000), etc.

**Figure 5.**
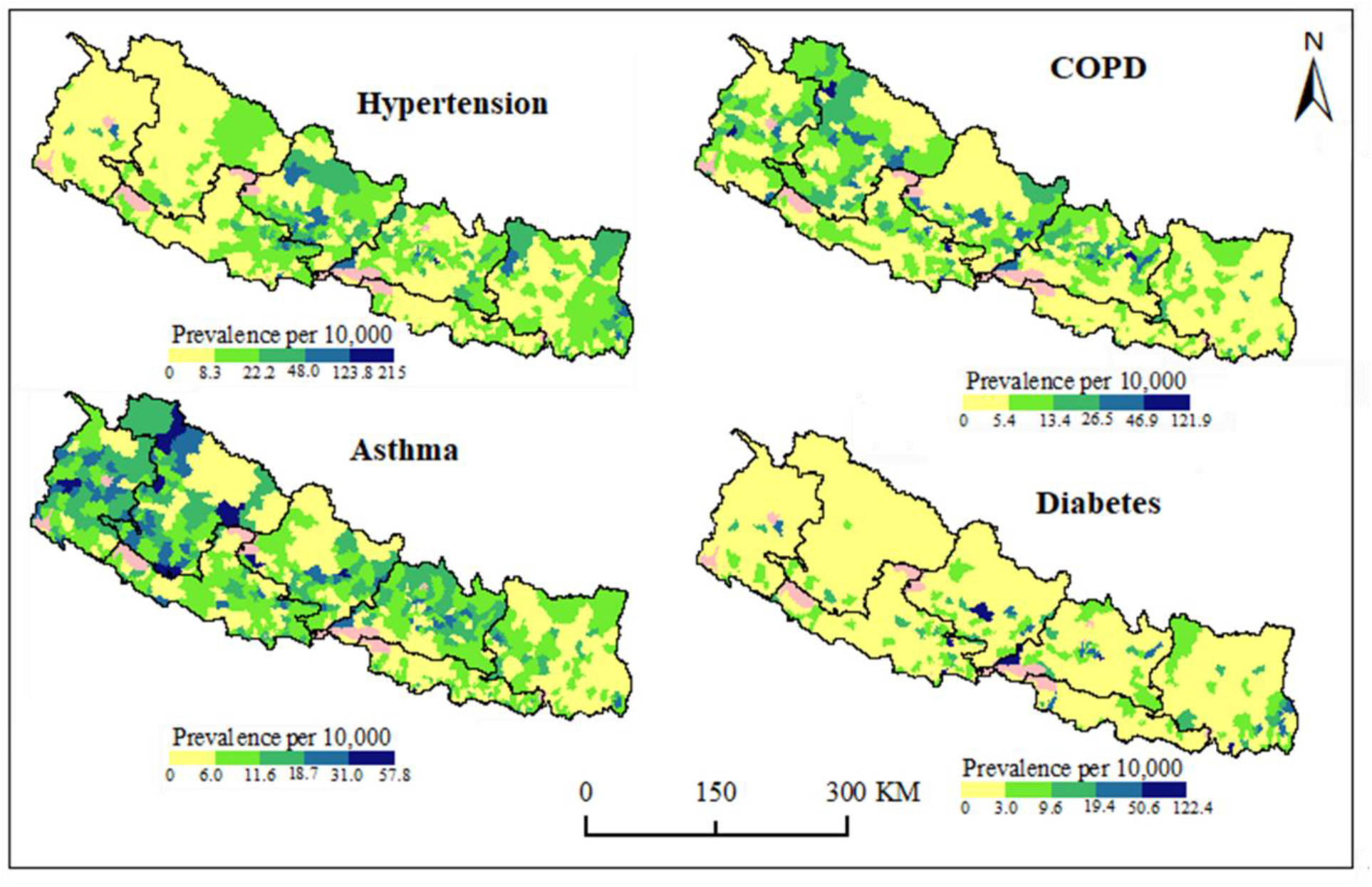
Spatial distribution of disease prevalence inequalities among the municipal units of Nepal

### 3.4 Vulnerability indices to the disease

The spatial assemblage of the social vulnerability index (SVI) was heterogeneous across 753 municipal units of Nepal (Fig. 6). It revealed that 44 (5.8%) municipal units (six municipalities and 38 rural municipalities) have very high vulnerability, however, population wise, 38.57% (10.7 million) of the total population of Nepal falls under highly vulnerable category (Table 2). Municipal units mainly in highland areas of eastern Nepal from Taplejung, Solukhumbu, Sankhuwasabha etc., from western Nepal such as Rukum, Dailekh, Bajura, Jajarkot, Dolpa etc. and Saptari district of Province No. 2 are among the highly vulnerable. The least vulnerable municipal units (149/753) are mainly located in better developed areas such as Jhapa, Morang, Sunsari, Kathmandu valley, Chitwan, Rupandehi, Dang, Kailali etc. Four metropolitan cities-Biratnagar, Lalitpur, Kathmandu and Bharatpur; three sub-metropolitan cities – Dharan, Itahari and Kalaiya; and 66 municiplaities come under the category of very low SVI.

**Table 2.**
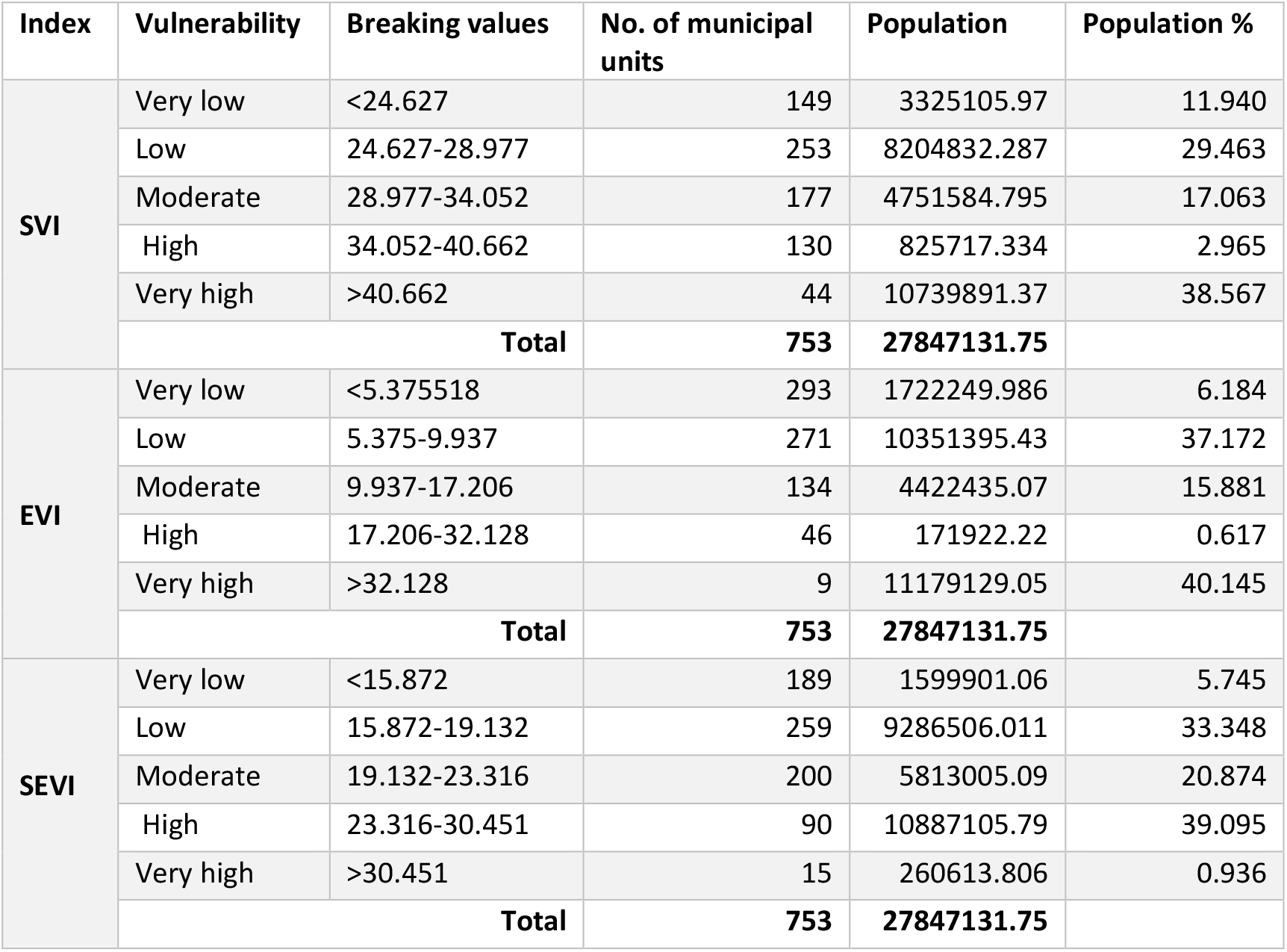
Spatial and demographic distribution of different categories of the vulnerability indices in Nepal

**Figure 6.**
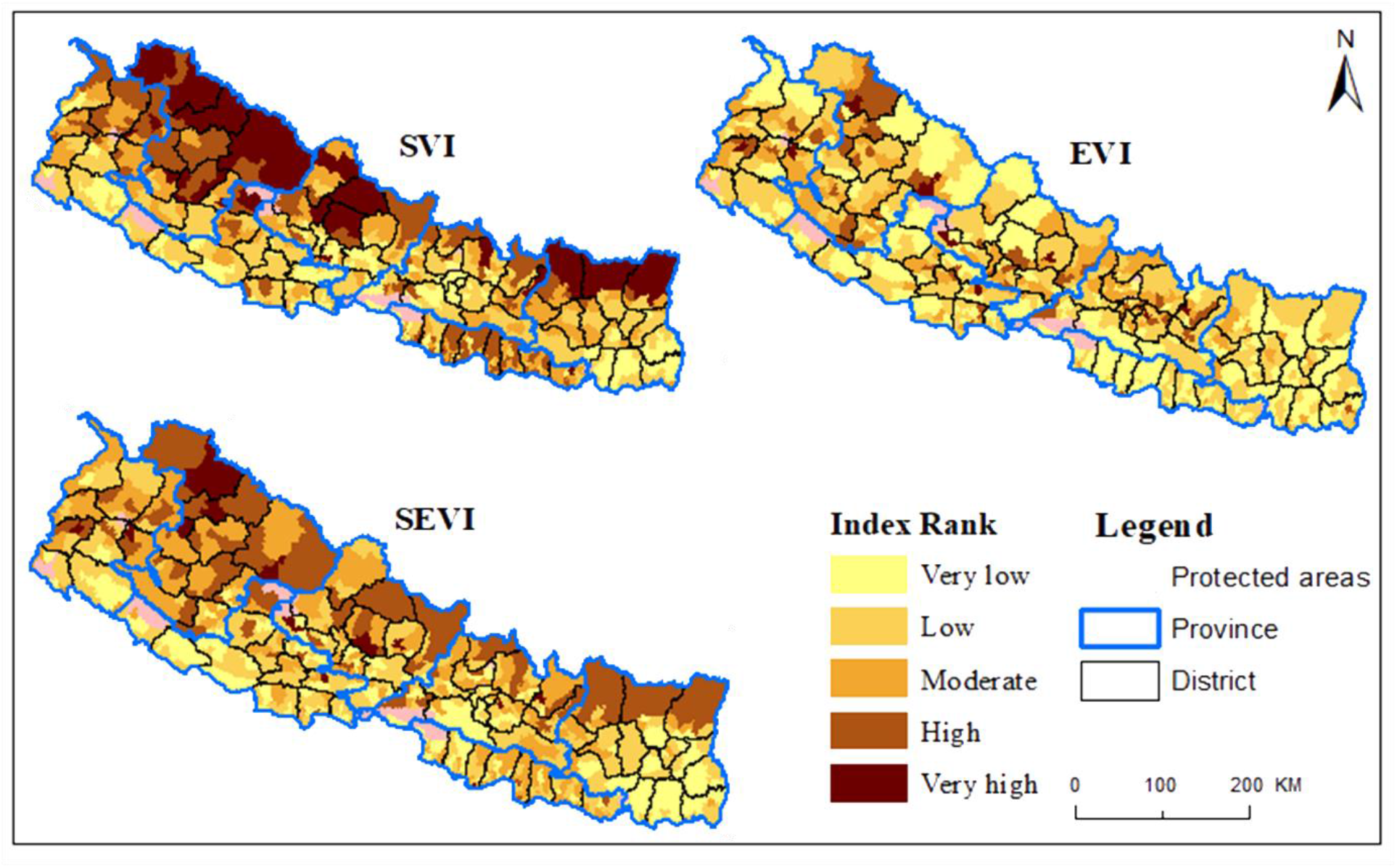
Geospatial arrangement of social vulnerability index (SVI), epidemiological vulnerability index (EVI) and social-epidemiological vulnerability index (SEVI), across 753 municipal units of Nepal grouped into five ranks.

There was no distinct pattern on epidemiological vulnerability index (EVI) among the municipal units of Nepal. The EVI identified nine municipal units as highly vulnerable, however, demographically, 40.15% (11.18 million) of the country population came under this category. Municipal units having high population density with higher prevalence of diseases like asthma, diabtetes, COPD and hypertension such as Dhulikhel, Tansen, Beshishahar, Amarghadhi municipalities fell under the class of very high epidemiological vulnerability index. Among five categories of the EVI, the highest number of municipal units (n= 293) fell under the ‘very low’ but only 6.18% of the total population belonged to this group.

The Social-Epidemiological Vulnerability Index (SEVI), a composite of the two (SVI and EVI) revealed that 15 municipal units of Nepal are under very high vulnerability of the disease. It included Pokhara Lekhnath metropolitan city, 10 municipalities (Besishahar, Thuli Bheri, Chhayanath Rara, Bhimeshwor, Dhulikhel, Gaur, Dhorpatan, Tansen, Sanphebagar and Amargadhi) and four rural municipalities from Karnali province (Palata, Chankheli, Kharpunath and Sarkegad). The SEVI revealed high vulnerability of 90 municipal units of Nepal including Bharatpur metropolitan city, 28 municipalities more abundantly from Karnali and Sudoorpashchim provinces; and 61 rural municipalities from all over Nepal. Demographically, about 40% (11.15 million) population (0.94% very high vulnerability and 39.1% high vulnerability) is highly vulnerable to epidemics. A total of 189 municipal units fall under the category of ‘very low’ SEVI, which include Lalitpur metropolitan city, five sub-metropolitan cities (Dharan, Itahari, Hetauda, Kalaiya and Tulsipur), 70 municipalities and 113 rural municipalities.

## 4 Discussion

The corona virus disease 19 (COVID-19) has become a global pandemic and lacks the vaccine and antiviral drug till date (25^th^ June 2020). For containing the disease within the lesser severity, it is important to know the level of vulnerability among the geographical units and the communities such that disease control interventions could be efficiently deployed to the highly vulnerable groups. Here, we present community-level vulnerability index to the epidemics for 753 municipal units under 77 districts of 7 provinces in Nepal. The results could be valuable for the local government to effectively control the transmission dynamics of the COVID-19 pandemic and other epidemics. Index based vulnerability assessment has been used in a number of diseases including COVID-19 (Dickin et al. 2013, DeCapprio et al. 2020, Macharia et al. 2020, KC et al. 2020). The high vulnerability score can be described as a location where a higher percentage of COVID-19 cases would result in severe outcomes such as hospitalization or death as compared to a locality with a low vulnerability score.

The social vulnerability index (SVI) varied significantly among the municipalities and rural municipalities of Nepal as a result of a spatial variation of the underlying vulnerability indicators. The SVI identified the municipal units as highly vulnerable ones that have higher population density, greater proportion of the elderly population, larger illiteracy rate and longer travel time to the hospitals and cities. The rate of disease spread increases with human population density and it is consistent for vector borne, air borne as well as contact transmission diseases. Patterns of illness and death due to COVID-19 reflect urban social and economic geographies because poor people living in the high densities with low quality housing seem more affected (Ahmed et al. 2020, Rocklöv & Sjödin 2020, Simon 2020). There is higher probability of interpersonal contact due to overcrowding, difficulties in maintaining the social distance and contact tracing of the victims etc. Greater proportion of the elderly population can be another factor that enhances the severity of the disease in a community. The COVID-19 has been observed to be more fatal to the people of age group above 60 who are already having compromised immunity due to pulmonary or/and cardiovascular disorders (Chau et al. 2014, Fang et al. 2020, Kassir 2020). Therefore, communities with high population density and higher proportion of elderly people are more vulnerable to the epidemics.

Inaccessibility factors such as travel time to the cities and hospitals are other important determinants of higher vulnerability to diseases. Lack of access to health care systems due to poverty and need of longer travel time to the health centers greatly compromises the adaptive capacity of communities against epidemics (Olago et al. 2007, Kienberger & Hagenlocher 2014). In line to these, our results revealed that municipal units across the entire higher mountain region of Nepal have higher vulnerability. The ability to prevent, detect, and respond to epidemics demand access to rapid and high-quality healthcare. Travel time negatively impacts healthcare-seeking behavior of local inhabitants (Ji et al. 2020, Rader et al. 2020). Rural municipalities across the mountain region are sparsely populated; therefore, chances of human-to-human transmission might be low. However, if the disease spreads, it can have high fatality due to inaccessibility to the health facilities to the illiterate and poor people therein.

Densely populated cities across Nepal and the districts of Province No. 2 are highly vulnerable. Higher level of pollution, lack of proper sanitation, illiteracy, overcrowding of people leading to failure in sustaining the interpersonal distance, lower public health related awareness among the people in those areas make the communities highly susceptible to epidemics. Improper waste disposal, unhygienic attitudes, deplorable sanitary conditions, poor knowledge of disease transmission mechanisms, unorthodox beliefs about epidemics, and ignorance on issues of health and healthy living contribute in higher severity of the epidemic (Ndah & Ngoran 2015). All eight districts of Province No 2 have much higher population density than rest of the districts of Nepal and more than half of the households lack toilet facilities.

The epidemiological vulnerability index (EVI) revealed that municipalities and rural municipalities that have higher poverty, lack of sanitation and population with higher frequency of respiratory and cardiovascular diseases (such as Dhulikhel, Tansen, Beshishahar, Sanphebagar, Amargadhi, etc.) are highly vulnerable to the epidemics. Urbanization and population density are not the sole causes of high COVID-19 infection rate, suburban areas with limited health and sanitation facilities are equally prone to the disease (Miller 2020). The rapidly urbanizing cities often bear fast growing unplanned settlements and lack basic infrastructure (Kienberger & Hagenlocher 2014), hence, suburban areas or growing cities become more vulnerable to epidemics. Among the eight municipal units belonging to the very high vulnerability group on EVI, seven are rapidly urbanizing municipalities (Beshishahar, Thulibheri, Bhimeswor, Dhulikhel, Dhorpatan, Tansen, Sanphebagar and Amargadhi in decreasing order of severity) and one (Sarkegad RM in Humla district) is rural municipality. Further, data revealed that all of those municipal units have very high prevalence of either all or some of the diseases like asthma, diabetes, COPD and hypertension. Elderly people with comorbidities especially those with hypertension, pulmonary disorders, coronary heart disease or diabetes are more likely to be infected and account for a large proportion of deaths from COVID-19 (Fang et al. 2020, Yang et al. 2020, Zheng et al. 2020).

The SEVI gave more smoothed results because it is derived as the average of the SVI and EVI. It accounted all the demographic, socioeconomic and public health related indicators and predicted almost 40% of the population under high vulnerability group. The areas identified as the highly vulnerable to the epidemics are densely populated cities or growing cities with under developed infrastructure, higher prevalence of pulmonary and cardio-vascular disorders and inaccessibility to the health facilities. The communities having higher proportion of ethnic minorities living in poverty with low levels of educational attainment have higher rates of hospitalization and death related to COVID-19 than the others (Platt & Warwick 2020, Wadhera et al. 2020). As the SEVI has been obtained by combined processing of demographic, social and epidemiological indicators all together, the vulnerability results could be applicable to other epidemics too.

Most of the confirmed cases of COVID-19 in Nepal till date are among the travelers who returned back from abroad including India. Community level transmission has been observed only in a few pocket areas such as Birgunj, Udaypur, Nepalgunj etc. (Tharu et al. 2020). Early isolation of the people by means of the forcefully imposed lockdown, awareness programs and terror of disease among people helped in breaking the transmission chain. However, as the number of cases is increasing exponentially, although limited among the immigrants so far, there is a high risk of community transmission from asymptomatic carriers of the SARS-CoV-2. Currently, in the absence of good enough representative cases of COVID-19 community transmission, we couldn’t test efficacies of the computed vulnerability indices. We believe that the vulnerability indices obtained by analyzing the demographic, socioeconomic and epidemiological indicators are valuable tools to the local and state governments for prioritization and improved planning especially in highly vulnerable communities where community level transmission can happen at any moment.

The quality of this assessment depends on the accuracy and completeness of the data used here. Specially, we suspect the completeness of epidemiological indicators as this information were derived from the OPD services. In addition, there might be other several factors needed to be considered in the commutation of COVID-19 related vulnerability index. For example, in the absence of organized data, we could not analyze the huge number of populations who are returning from COVID-19 affected countries like India, middle east Asian countries and Malaysia.

## 5 Conclusions

We observed heterogeneity in the distribution of the community vulnerability to epidemics among the 753 municipalities and rural municipalities of Nepal. Most of the municipal units from Province No 2, Karnali and Sudoorpashchim provinces are highly vulnerable to the COVID-19. Additionally, some fast-growing cities with higher prevalence of pulmonary and cardiovascular disorders, lack of proper sanitation are vulnerable to the disease. Longer travel time required to access the health facilities makes the municipal units of mountain region more vulnerable than the rest. Such nature of the vulnerability emphasizes the need to address social determinants of health discrepancies and formulate management programs to deploy the resources efficiently to highly vulnerable communities.

## Data Availability

All data used are freely available in the depositories that are clearly mentioned in the methods section.

## Acknowledgement

We would like to thank Dr. Meghnath Dhimal, Nepal Health Research Council (NHRC) for his support. We acknowledge sources of data particularly, Health Management Information System (HMIS), Government of Nepal, Demographic and Health Survey (DHS) and Worldpop.

## Author contribution

LK and BKA conceptualized the study. LK, BKA, BP gathered the data. LK and BKA analyzed the data and wrote the manuscript. All authors contributed in the manuscript improvement and approved it for the submission.

## References

Acharya, B. K., Cao, C., Lakes, T., Chen, W., Naeem, S. and Pandit, S. 2018. Modeling the spatially varying risk factors of dengue fever in Jhapa district, Nepal, using the semi-parametric geographically weighted regression model. International Journal of Biometeorology 62:1973–1986. https://doi.org/10.1007/s00484-018-1601-8.

Adger, W. N. 2006. Vulnerability. Global Environmental Change 16:268–281. https://doi.org/10.1016/j.gloenvcha.2006.02.006.

Ahmed, F., Ahmed, N. e., Pissarides, C. and Stiglitz, J. 2020. Why inequality could spread COVID-19. The Lancet Public Health 5:e240. https://doi.org/10.1016/s2468-2667(20)30085-2.

Anderson, R. M., Heesterbeek, H., Klinkenberg, D. and Hollingsworth, T. D. 2020. How will country-based mitigation measures influence the course of the COVID-19 epidemic? The Lancet 395:931–934. https://doi.org/10.1016/s0140-6736(20)30567-5.

Anderson, R. M., Heesterbeek, H., Klinkenberg, D. and Hollingsworth, T. D. 2020. How will country-based mitigation measures influence the course of the COVID-19 epidemic? The Lancet 395:931–934. https://doi.org/10.1016/S0140-6736(20)30567-5.

Anselin, L., Syabri, I. and Kho, Y. 2006. GeoDa: An introduction to spatial data analysis. Geographical Analysis 38:5–22.

Bates, I., Fenton, C., Gruber, J., Lalloo, D., Lara, A. M., Squire, S. B., et al. 2004. Vulnerability to malaria, tuberculosis, and HIV/AIDS infection and disease. Part II: determinants operating at environmental and institutional level. The Lancet Infectious Diseases 4:368–375. https://doi.org/10.1016/s1473-3099(04)01047-3.

Baud, D., Qi, X., Nielsen-Saines, K., Musso, D., Pomar, L. and Favre, G. 2020. Real estimates of mortality following COVID-19 infection. The Lancet Infectious Diseases. https://doi.org/10.1016/s1473-3099(20)30195-x.

Boe, D. M., Boule, L. A. and Kovacs, E. J. 2017. Innate immune responses in the ageing lung. Clinical and Experimental Immunology 187:16–25. https://doi.org/10.1111/cei.12881.

Cannon, T. 1994. Vulnerability analysis and the explanation of ‘Natural’ disasters. In: A. Varley (Eds) Disasters, Development and Environment. Wiley London: 13–30.

Chau, P. H., Gusmano, M. K., Cheng, J. O., Cheung, S. H. and Woo, J. 2014. Social vulnerability index for the older people-Hong Kong and New York City as examples. Journal of Urban Health 91:1048–1064. https://doi.org/10.1007/s11524-014-9901-8.

Coccia, M. 2020. Factors determining the diffusion of COVID-19 and suggested strategy to prevent future accelerated viral infectivity similar to COVID. Science of the Total Environment 729:138474. https://doi.org/10.1016/j.scitotenv.2020.138474.

DeCapprio, D., Burgess, T., Gartner, J., McCall, C. J., Sayed, S. and Kothari, S. 2020. Building a COVID-19 Vulnerability Index [Preprint]. 2003.07347.

Dickin, S. K., Schuster-Wallace, C. J. and Elliott, S. J. 2013. Developing a vulnerability mapping methodology: applying the water-associated disease index to dengue in Malaysia. PLoS One 8:e63584. https://doi.org/10.1371/journal.pone.0063584.

Dong, Y., Mo, X., Hu, Y., Qi, X., Jiang, F., Jiang, Z., et al. 2020. Epidemiology of COVID-19 among children in China. Pediatrics 145:e20200702. https://doi.org/10.1542/peds.2020-0702.

Fang, L., Karakiulakis, G. and Roth, M. 2020. Are patients with hypertension and diabetes mellitus at increased risk for COVID-19 infection? The Lancet Respiratory Medicine 8:e21. https://doi.org/10.1016/s2213-2600(20)30116-8.

Frankel, L. K. 2011. The relation of life insurance to public hygiene. 1910. American Journal of Public Health 101:1868–1869. https://doi.org/10.2105/ajph.2011.101101868.

Huang, C., Wang, Y., Li, X., Ren, L., Zhao, J., Hu, Y., et al. 2020. Clinical features of patients infected with 2019 novel coronavirus in Wuhan, China. The Lancet 395:497–506. https://doi.org/10.1016/s0140-6736(20)30183-5.

Jenks, G. 1977. Optimal data classification for choropleth maps Occasional (No. 2). University 152 of Kansas, Department of Geography.

Ji, Y., Ma, Z., Peppelenbosch, M. P. and Pan, Q. 2020. Potential association between COVID-19 mortality and health-care resource availability. The Lancet Global Health 8:e480. https://doi.org/10.1016/s2214-109x(20)30068-1.

Kamel Boulos, M.N. and Geraghty, E. M. 2020. Geographical tracking and mapping of coronavirus disease COVID-19/severe acute respiratory syndrome coronavirus 2 (SARS-CoV-2) epidemic and associated events around the world: how 21st century GIS technologies are supporting the global fight against outbreaks and epidemics. International Journal of Health Geography 19:8. https://doi.org/10.1186/s12942-020-00202-8.

Kang, D., Choi, H., Kim, J. H. and Choi, J. 2020. Spatial epidemic dynamics of the COVID-19 outbreak in China. International Journal of Infectious Diseases 94:96–102. https://doi.org/10.1016/j.ijid.2020.03.076.

Kassir, R. 2020. Risk of COVID-19 for patients with obesity. Obesity Reviwes 21:e13034. https://doi.org/10.1111/obr.13034.

Kc, S., Mishra, R., Mishra, R. and Shukla, A. 2020. Community COVID-19 vulnerability index in India. Laxenburg, Austria, IIASA Working Paper.

Kienberger, S. and Hagenlocher, M. 2014. Spatial-explicit modeling of social vulnerability to malaria in East Africa. International Journal of Health Geographics 13:29.

Liu, Y., Gayle, A. A., Wilder-Smith, A. and Rocklov, J. 2020. The reproductive number of COVID-19 is higher compared to SARS coronavirus. Journal of Travel Medicine 27. https://doi.org/10.1093/jtm/taaa021.

Macharia, P. M., Joseph, N. K. and Okiro, E. A. 2020. A vulnerability index for COVID-19: spatial analysis to inform equitable response in Kenya [Preprint]. medRxiv. https://doi.org/10.1101/2020.05.27.20113803.

Mayala, B. K., Trinadh Dontamsetti, T. D. F. and C., T. N. 2019. Interpolation of DHS Survey Data at Subnational Administrative Level 2. DHS Spatial Analysis Reports No. 17.

Miller, J. 2020. The overstated COVID-19 blame on urban density in favor of suburban living. Forbes, 14th May 2020. https://www.forbes.com/sites.

MoHP, G. o. N. 2020. Ministry of health and popultion, government of Nepal. Retrieved 18th June 2020, from https://covid19.mohp.gov.np/#/.

Mollalo, A., Vahedi, B. and Rivera, K. M. 2020. GIS-based spatial modeling of COVID-19 incidence rate in the continental United States. Science of the Total Environment 728:138884. https://doi.org/10.1016/j.scitotenv.2020.138884.

Mwarumba, N. 2017. Global social vulnerability to pandemics: An examinationn of social determinants of H1N1 2009 mortality. PhD Thesis, Oklahoma State University, Oklahoma. Pp:150.

Ndah, A. B. and Ngoran, S. D. 2015. Liaising Water Resources Consumption, Urban Sanitation and Cholera Epidemics in Douala, Cameroon: A Community Vulnerability Assessment. Journal of Resources Development and Management 8:63–78.

Olago, D., Marshall, M., Wandiga, S. O., Opondo, M., Yanda, P. Z., Kangalawe, R., et al. 2007. Assessment of the potential of ecolabels to promote agrobiodiversity. Ambio 36:551–558. https://doi.org/10.1579/0044-7447(2007)36[551:aotpoe]2.0.co;2.

Platt, L. and Warwick, R. 2020. Are some ethnic groups more vulnerable to COVID-19 than others? The Institute for Fiscal Studies, Nuffield Foundation.

Pringle, D. G. 1996. Mapping disease risk estimates based on small numbers: An assessment of empirical Bayes techniques. Economic & Social Review 27:341–363.

Rader, B., Astley, C. M., Sy, K. T. L., Sewalk, K., Hswen, Y., Brownstein, J. S., et al. 2020. Geographic access to United States SARS-CoV-2 testing sites highlights healthcare disparities and may bias transmission estimates. Journal of Travel Medicine. https://doi.org/10.1093/jtm/taaa076.

Rader, B., Astley, C. M., Sy, K. T. L., Sewalk, K., Hswen, Y., Brownstein, J. S., et al. 2020. Increased travel times to United States SARS-CoV-2 testing sites: A spatial modeling study [Preprint]. Epidemiology. https://doi.org/10.1101/2020.04.25.20074419.

Ray, N. and Ebener, S. 2008. AccessMod 3.0: computing geographic coverage and accessibility to health care services using anisotropic movement of patients. International Journal of Health Geographics 7:63. https://doi.org/10.1186/1476-072X-7-63.

Rocklöv, J. and Sjödin, H. 2020. High population densities catalyse the spread of COVID-19. Journal of Travel Medicine 27:taaa038. https://doi.org/10.1093/jtm/taaa038.

Rothan, H. A. and Byrareddy, S. N. 2020. The epidemiology and pathogenesis of coronavirus disease (COVID-19) outbreak. Journal of Autoimmunity 109:102433. https://doi.org/10.1016/j.jaut.2020.102433.

Sarwar, S., Waheed, R., Sarwar, S. and Khan, A. 2020. COVID-19 challenges to Pakistan: Is GIS analysis useful to draw solutions? Science of the Total Environment 730:139089. https://doi.org/10.1016/j.scitotenv.2020.139089.

Semenza, J. C. and Giesecke, J. 2008. Intervening to reduce inequalities in infections in Europe. American Journal of Public Health 98:787–792. https://doi.org/10.2105/AJPH.2007.120329).

Simon, D. 2020. Cities are at centre of coronavirus pandemic - understanding this can help build a sustainable, equal future. The Conversation, 23 April 2020. https://theconversation.com.

Smith, J. A. and Judd, J. 2020. COVID-19: Vulnerability and the power of privilege in a pandemic. Health Promotion Journal of Australia 31:158–160. https://doi.org/10.1002/hpja.333.

Sominsky, L., Walker, D. W. and Spencer, S. J. 2020. One size does not fit all - Patterns of vulnerability and resilience in the COVID-19 pandemic and why heterogeneity of disease matters. Brain, Behavior, and Immunity. https://doi.org/10.1016/j.bbi.2020.03.016.

Sullivan, C. A. and Meigh, J. 2006. Integration of the biophysical and social sciences using an indicator approach: Addressing water problems at different scales. Water Resources Management 21:111–128. https://doi.org/10.1007/s11269-006-9044-0.

Tharu, T., Gahatraj, R., Shahi, M. and Gautam, G. 2020. Community transmission of COVID-19 in Nepalgunj city, Nepal (in Nepali language). Kantipur National Daily, 4th May 2020 http://www.ekantipur.com.

Tuite, A., Ng, V., Rees, E. and Fisman, D. 2020. Estimation of COVID-19 outbreak size in Italy based on international case exportations [Preprint]. medRxiv 20030049. https://doi.org/10.1101/2020.03.02.20030049.

Uddin, K., Shrestha, H. L., Murthy, M. S., Bajracharya, B., Shrestha, B., Gilani, H., et al. 2015. Development of 2010 national land cover database for the Nepal. Journal of Environmental Management 148:82–90. https://doi.org/10.1016/j.jenvman.2014.07.047.

Wadhera, R. K., PriyaWadhera, Gaba P., Figueroa, J. F., Maddox, K. E. J., Yeh, R., et al. 2020. Variation in COVID-19 hospitalizations and deaths across New York city boroughs. JAMA 323:2194–2195. https://doi.org/10.1001/jama.2020.6887.

Weiss, D. J., Nelson, A., Gibson, H. S., Temperley, W., Peedell, S., Lieber, A., et al. 2018. A global map of travel time to cities to assess inequalities in accessibility in 2015. Nature 553:333–336. https://doi.org/10.1038/nature25181.

WHO. 2020a. Report of the World Health Organization-China Joint Mission on Coronavirus Disease 2019 (COVID-19). Retreived from: https://www.who.int/docs/default-source/coronaviruse/who-china-joint-mission-on-covid-19-final-report.pdf.

WHO. 2020b. Coronavirus Disease 2019 (COVID-19) Situation Report - 157. World Health Organization. Retrieved from: https://www.who.int/docs/default-source/coronaviruse/situation-reports/20200625-covid-19-sitrep-157.pdf?sfvrsn=423f4a82_2.

Wu, Z. and McGoogan, J. M. 2020. Characteristics of and important lessons from the coronavirus disease 2019 (COVID-19) outbreak in China summary of a report of 72314 cases from the Chinese Center for Disease Control and Prevention. JAMA 323:1239–1242. https://doi.org/10.1001/jama.2020.2648.

Yang, Y., Peng, F., Wang, R., Guan, K., Jiang, T., Xu, G., et al. 2020. The deadly coronaviruses: The 2003 SARS pandemic and the 2020 novel coronavirus epidemic in China. Journal of Autoimmunity 109:102434. https://doi.org/10.1016/j.jaut.2020.102434.

You, D., Lindt, N., Allen, R., Hansen, C., Beise, J. and Blume, S. 2020. Migrant and displaced children in the age of COVID-19: How the pandemic is impacting them and what we can do to help. Migration Policy Practice X:32–39.

Zheng, Y. Y., Ma, Y. T., Zhang, J. Y. and Xie, X. 2020. COVID-19 and the cardiovascular system. Nature Reveiws Cardiology 17:259–260. https://doi.org/10.1038/s41569-020-0360-5.

